# Prototyping OSA: A New Framework for Predicting Cardiovascular Outcomes and CPAP Benefit

**DOI:** 10.64898/2026.10.30.26366770

**Authors:** Tahir Hafeez Malik, David Daniel, Jo Hsuan Lee, Zainab Al-Taie, Kavya Devarakonda, Samira Khan, Vi Le, Kelly A. Loffler, Craig S. Anderson, Mathias Baumert, Jason C. Kovacic, Shishir Adhikari, Vaishnavi Kundel, Cameron S. McAlpine, Girish Nadkarni, Oren Cohen, Mayte Suárez-Fariñas, Neomi Shah

## Abstract

**Rationale:** Obstructive sleep apnea (OSA) is associated with major adverse cardiac events (MACE); continuous positive airway pressure (CPAP) has not reduced MACE in randomized trials. We hypothesized that multidomain machine learning would identify OSA subgroups differing in cardiovascular risk and CPAP response.

**Methods:** We conducted a secondary analysis of SAVE trial data. After preprocessing, Gaussian mixture models were fitted; Bayesian information criterion and C-index selected five prototypes. Cox models estimated MACE risk by prototype. Baseline quality of life (EQoL), Epworth Sleepiness Scale (ESS), and Framingham Risk Score (FRS) were compared; an existing causal survival forest model estimated prototype-specific CPAP effects on cardiovascular risk.

**Measurements and Main Results:** After preprocessing, five OSA prototypes were identified among 2,676 participants using 88 baseline variables. Prototypes differed in selected demographic, cardiometabolic, medication, and sleep variables, including smoking status, diabetes, prior myocardial infarction/stroke, antiplatelet/lipid-lowering therapy, BMI, oxygenation metrics, apnea-hypopnea index, and ESS (overall p<0.001). They also differed in baseline EQoL, FRS, MACE rate, and predicted CPAP response. Prototype 1 had the highest ESS, FRS, and MACE rate, whereas Prototype 5 had the lowest FRS and MACE rate. Using the causal survival forest model to assess prototype-specific CPAP effects on cardiovascular risk, Prototype 5 was predicted to benefit from CPAP (p<0.05), whereas Prototypes 1, 3, and 4 showed no benefit or possible harm.

**Conclusion:** These findings demonstrate heterogeneity in cardiovascular risk and predicted CPAP response, potentially explaining neutral trial results. After external validation, these prototypes could guide trial enrichment and targeted CPAP use to improve cardiovascular outcomes.

**Impact Statement:** This study advances clinical medicine by identifying multidomain OSA prototypes that differ in baseline cardiovascular risk and predicted cardiovascular response to CPAP, potentially supporting more precise treatment selection and future cardiovascular-outcomes trials. It adds to the disease knowledge base by showing that OSA-related cardiovascular risk is not captured by conventional severity metrics alone, but reflects interactions among sleep, cardiometabolic, demographic, and clinical characteristics. These findings provide a framework for mechanistic and prospective studies of the biological pathways underlying heterogeneous cardiovascular susceptibility and CPAP response in OSA.

## Introduction

Obstructive sleep apnea (OSA) is characterized by repetitive upper airway collapse, resulting in intermittent hypoxemia and increased sympathetic tone, both of which are thought to contribute to cardiovascular disease (CVD) (1–4). OSA is estimated to affect nearly one billion individuals globally (5). Additionally, CVD remains the leading cause of morbidity and mortality worldwide (CITE), and prevention increasingly depends on identifying and modifying upstream risk factors. However, randomized controlled trials, including Sleep Apnea Cardiovascular Endpoints (SAVE), Randomized Intervention With Continuous Positive Airway Pressure (CPAP) in Coronary Artery Disease and Sleep Apnea (RICCADSA), and Impact of Sleep Apnea Syndrome in the Evolution of Acute Coronary Syndrome (ISAACC) have shown no significant reduction in composite CVD events among patients randomized to CPAP, compared with usual care (6–8). These consistent yet surprising findings raise the critical question of whether there is heterogeneity in the OSA population and whether a “one-size-fits-all” approach to treatment for all OSA subgroups with CPAP obscures differential cardiovascular risk profiles and treatment responses.

Several recent studies suggest that OSA is not a uniform disorder but rather a heterogeneous syndrome with distinct subgroups. For example, several studies have identified symptom-based OSA subtypes associated with differential cardiovascular outcomes (9–11). Other work has identified physiologic biomarkers such as hypoxic burden and heart-rate response to respiratory events that may predict adverse cardiovascular outcomes more robustly than the conventional OSA Apnea-Hypopnea Index (AHI) (12, 13). Although these studies advanced the field, they relied on limited data domains which do not capture the full complexity of OSA patient presentations. Analyzing the rich multidimensional data (including clinical histories, demographics, laboratory values, and polysomnography (PSG)) via machine learning is a next step in capturing the complexity within the OSA population (14, 15).

This study applies unsupervised clustering methods to data from the SAVE trial to identify biologically and clinically meaningful OSA prototypes using a comprehensive, data-driven approach rather than relying on individual metrics or predefined combinations of sleep metrics. We then determine whether these prototypes identify distinct patterns of cardiovascular risk, symptom burden, quality of life, and heterogeneous cardiovascular responses to CPAP therapy, including subgroups that may benefit from or be harmed by CPAP, advancing precision medicine in OSA.

## Methods

### Participants and Outcomes

We performed a secondary analysis of participants with OSA enrolled in the SAVE trial (ClinicalTrials.gov ID: NCT00738179). Detailed trial methods have been published previously (McEvoy et al., 2016). Briefly, SAVE enrolled patients with moderate-to-severe OSA, defined as an oxygen desaturation index (ODI) >12 events/hour, and established CVD from 89 centers across 7 countries. Patients with excessive daytime sleepiness, defined as an Epworth Sleepiness Scale (ESS) score >15, were excluded.

Baseline variables were recorded across nine data domains: substance use, comorbidities, general and mental health, demographics, cardiovascular vital signs, cardiovascular medication use, Apnealink Cardiorespiratory Polygraphy data, exercise, and sleep apnea signs. Participants were followed for the primary composite cardiovascular endpoint of death from cardiovascular causes, myocardial infarction, stroke, or hospitalization for unstable angina, heart failure, or transient ischemic attack. The full study design and protocol are detailed in the SAVE trial publication and its Supplementary Appendix (6).

Local institutional review board approval for this post hoc analysis was obtained from the Icahn School of Medicine at Mount Sinai in New York, New York (#21-01273).

### Data Pre-Processing

All analyses were performed in R, version 4.5.2. We evaluated 216 baseline variables from the nine baseline domains described above for inclusion in prototype development.

Variables judged by two independent clinician reviewers not to be routinely available in clinical care were removed; disagreements were adjudicated by a third clinician reviewer. Variables with >20% missingness were excluded to limit reliance on imputation, and categorical variables in which >95% of observations fell within a single category were excluded because of low variability. For retained categorical variables, sparse categories representing <5% of observations were reviewed and, when clinically appropriate, merged with related categories to reduce sparsity before imputation and modeling. Missing values in the remaining variables were imputed using additive linear regression with bootstrapping over 10 resamples, implemented with the Hmisc package in R.

After variable filtering and imputation, the processed variable dataset was reduced to a 10-dimensional representation using uniform manifold approximation and projection (UMAP) with Gower distance to account for mixed continuous and categorical data. A two-dimensional projection of this UMAP representation was used to visualize sample relationships and identify potential outliers. Samples with extreme separation in this two-dimensional projection, defined as >5 standard deviations from the cohort distribution, were classified as outliers and excluded from subsequent analyses. The retained 10-dimensional UMAP coordinates for the remaining 2,676 participants were used for clustering.

### Clustering

Gaussian mixture models (GMMs) were fitted to the 10-dimensional UMAP representation of the final analytic cohort to identify data-driven patient subgroups. For each clustering solution, GMMs estimated each participant’s posterior probability of membership in each subgroup, and participants were assigned to the subgroup for which they had the highest posterior probability. We evaluated solutions ranging from two to ten clusters and compared model fit using the Bayesian information criterion (BIC). The final clustering solution was selected using the C-index from the clusterCrit package, prioritizing within-cluster cohesion and between-cluster separation (16). The resulting clusters are hereafter referred to as OSA prototypes.

### Prototype Characterization and Feature Importance

After the final clustering solution was selected, we characterized each prototype by identifying variables that most strongly discriminated prototype membership. Variable-level importance was estimated using the Boruta algorithm, a random forest–based feature selection method.

Boruta evaluates variable importance by comparing each observed variable with shadow variables, defined as randomly permuted copies of the original cluster membership that represent the null distribution of importance. In each iteration, a random forest classifier was trained to predict prototype membership, and variable importance was quantified using mean decrease in accuracy, which reflects the reduction in classification accuracy after the relationship between a variable and the outcome is disrupted by permutation. Observed variables were compared with the distribution of shadow-variable importance scores across 1,000 iterations. Variables with Bonferroni-adjusted p values <0.01 were classified as confirmed predictors, whereas variables with importance scores significantly lower than the shadow distribution were rejected. Variables that remained unresolved after the maximum number of iterations were classified as tentative. Confirmed and tentative variables were retained for subsequent prototype characterization. Variable-level importance was visualized using violin plots showing the distribution of Boruta importance scores across the 1,000 iterations.

To summarize the relative contribution of broader clinical domains, confirmed and tentative variables were grouped according to their domains. Domain-level importance was estimated by aggregating the importance of variables within each domain using a mixed-effects modeling framework. This analysis was used to identify the clinical domains that contributed most strongly to prototype differentiation.

Finally, to support clinical interpretation of the prototypes, we summarized the distribution of retained variables across prototypes using heatmaps. For continuous variables, prototype-specific averages were standardized and displayed across prototypes. For categorical variables, prototype-specific proportions were standardized and displayed across prototypes. These heatmaps were used to visualize the clinical features that distinguished each prototype.

### Cluster Analysis – Outcomes

We compared baseline ESS scores across prototypes using pairwise Student’s t-tests. Baseline European Quality of Life (EQoL) utility scores and Framingham Risk Scores (FRS) were compared across prototypes using pairwise Mann–Whitney U tests. Time-to-event analyses for SAVE’s primary composite cardiovascular endpoint and its component events were performed using Cox-proportional hazards regression. Prototype-specific cardiovascular event risk was estimated relative to the overall SAVE cohort.

Because prototype-level treatment analyses estimate average CPAP effects within each subgroup, they may obscure heterogeneity in treatment response among individuals assigned to the same prototype. In addition, although treatment assignment was randomized in the parent SAVE trial, post hoc subgroup analyses may be limited by smaller sample sizes and incomplete covariate balance within data-derived subgroups; therefore, prototype-level treatment estimates may not fully preserve the causal advantages of the original randomized design.

To assess individualized treatment responsiveness, we applied a previously developed causal survival forest model (17), which estimates the individual treatment effect (ITE) of CPAP for each participant using baseline covariates. The model estimates each participant’s predicted cardiovascular risk had they been randomized to the alternative treatment allocation, thereby generating participant-level estimates of predicted CPAP benefit or harm. ITE estimates were aggregated within each prototype to assess prototype-specific predicted treatment responsiveness. Prototype-level mean ITE estimates and 95% confidence intervals were calculated to determine whether predicted CPAP effects differed from the null value of no treatment effect.

## Results

### Data Pre-Processing

A total of 2,687 SAVE participants formed the basis for analysis. Of 216 baseline variables evaluated, 84 were excluded after clinician review, 34 were excluded because of >20% missingness, and 10 categorical variables were excluded because >95% of observations fell within a single category (Table S1). For retained categorical variables, sparse categories representing <5% of observations were merged with clinically related categories when appropriate. After filtering and imputation, 88 variables were retained and projected into a 10-dimensional UMAP representation to preserve the structure and variability of the multidimensional baseline data. Eleven samples met the prespecified outlier criterion on two-dimensional UMAP visualization and were excluded from downstream analyses, yielding a final analytic cohort of 2,676 participants (Figure S1).

### Baseline Characteristics

After preprocessing and outlier exclusion, the final analytic cohort included 2,676 participants. Baseline characteristics are summarized in Table 1. Participants had a mean age of 61.3 ± 7.8 years, and 2,165 (80.9%) were male. Mean body mass index (BMI) was 28.6 ± 4.5 kg/m², mean apnea-hypopnea index (AHI) was 29.1 ± 16.0 events/hour, and mean ESS score was 7.4 ± 3.6. Current smoking was reported in 405 participants (15.2%), hypertension in 2,100 (78.7%), and diabetes mellitus in 797 (29.9%).

**Table 1.**
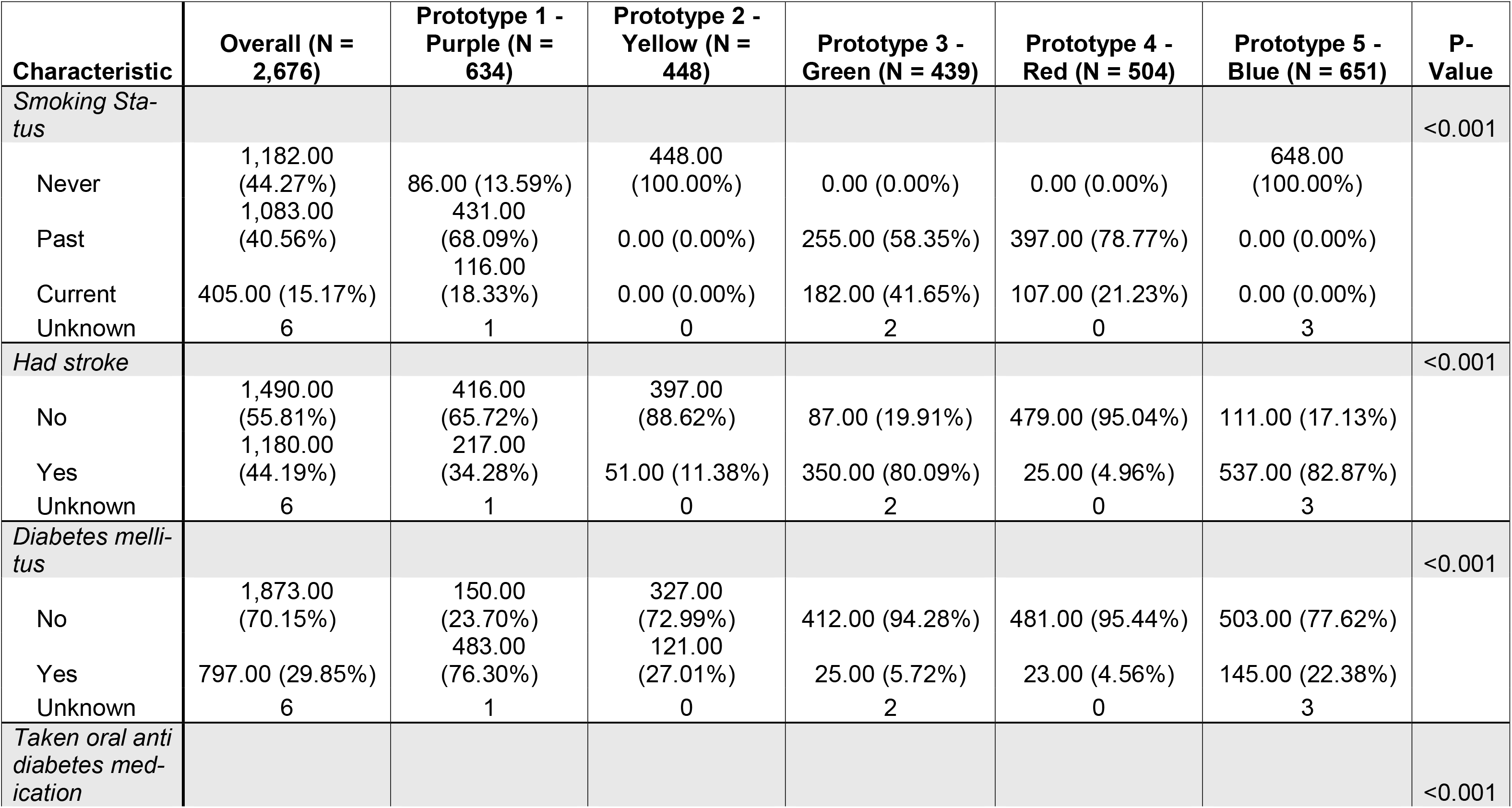
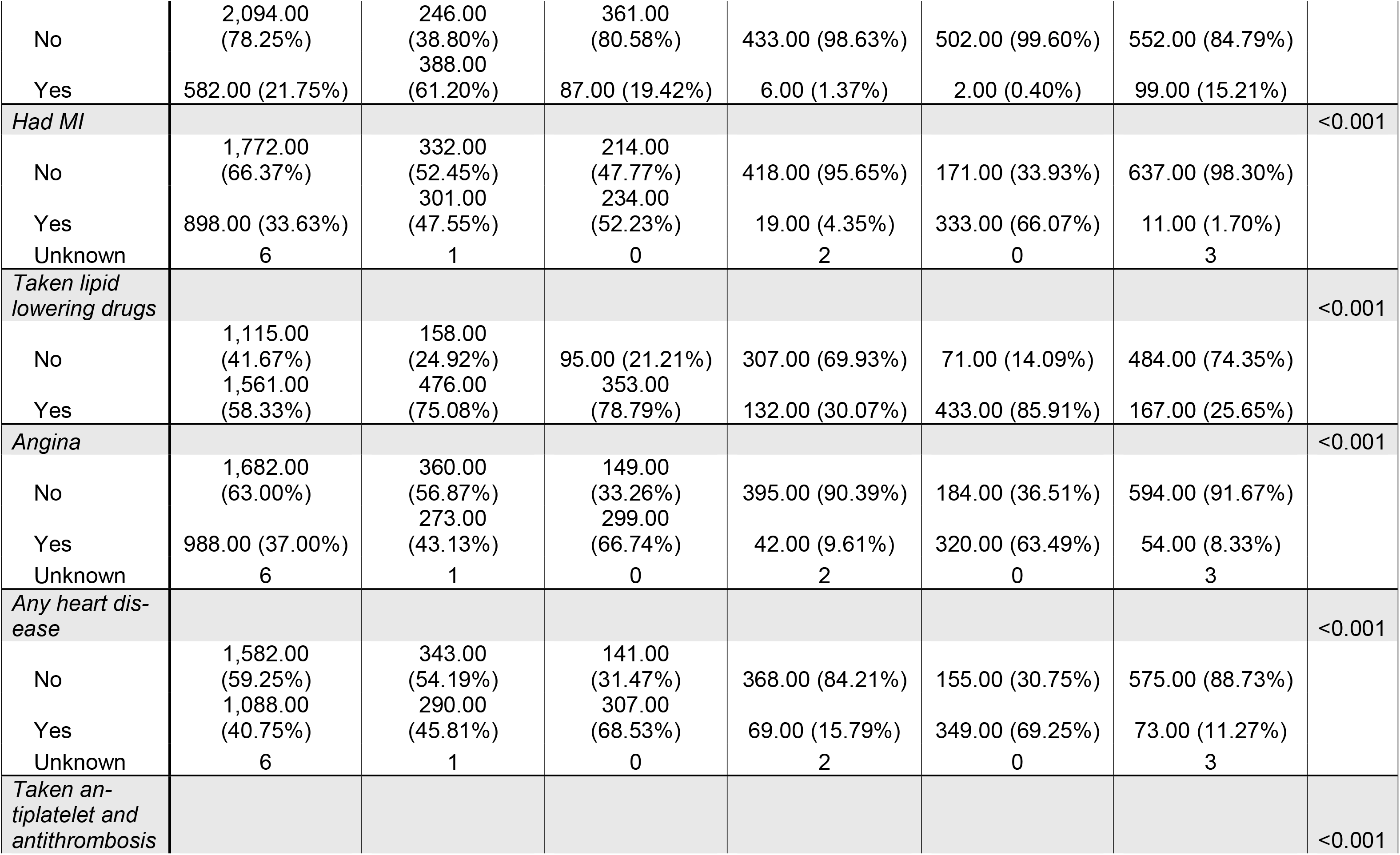
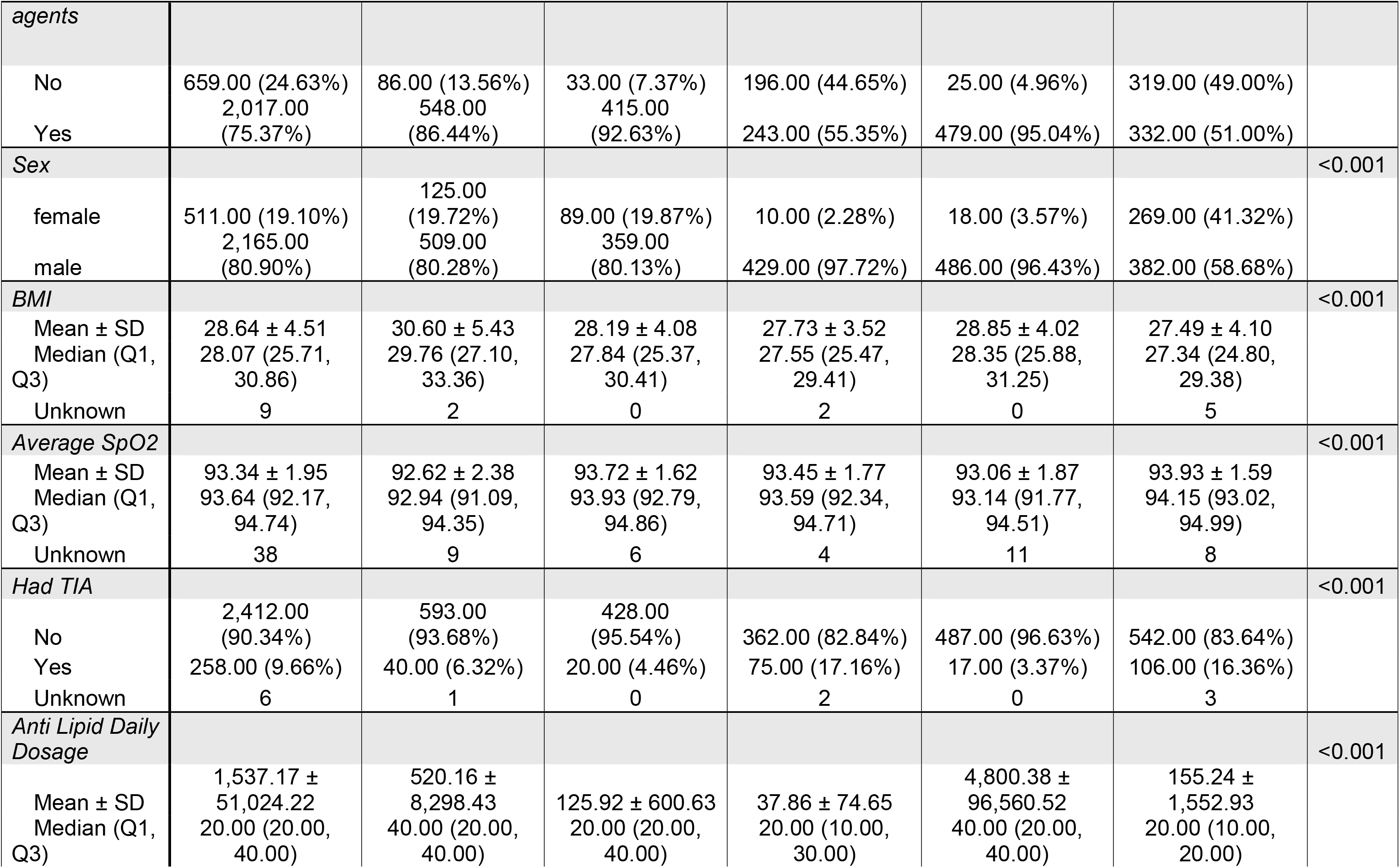
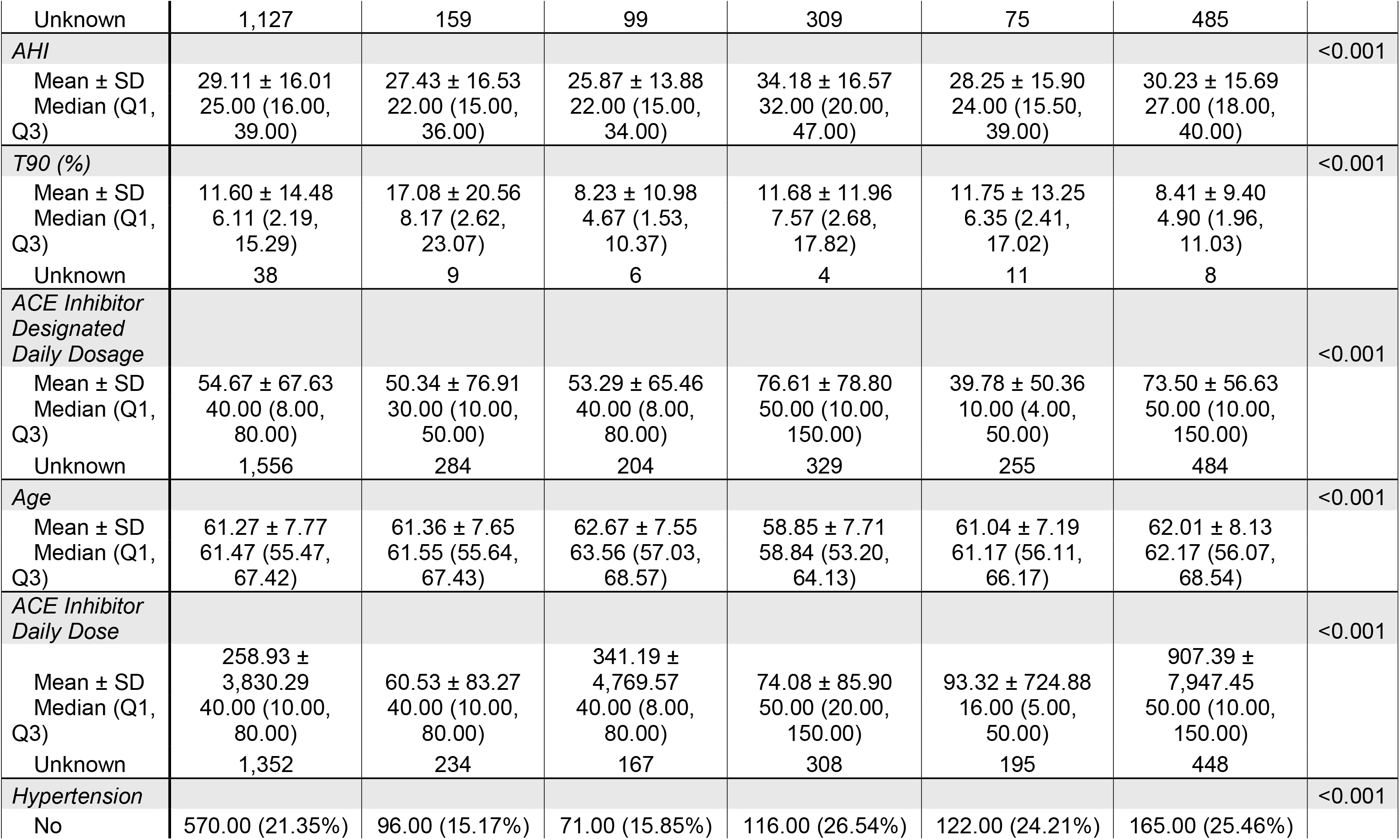
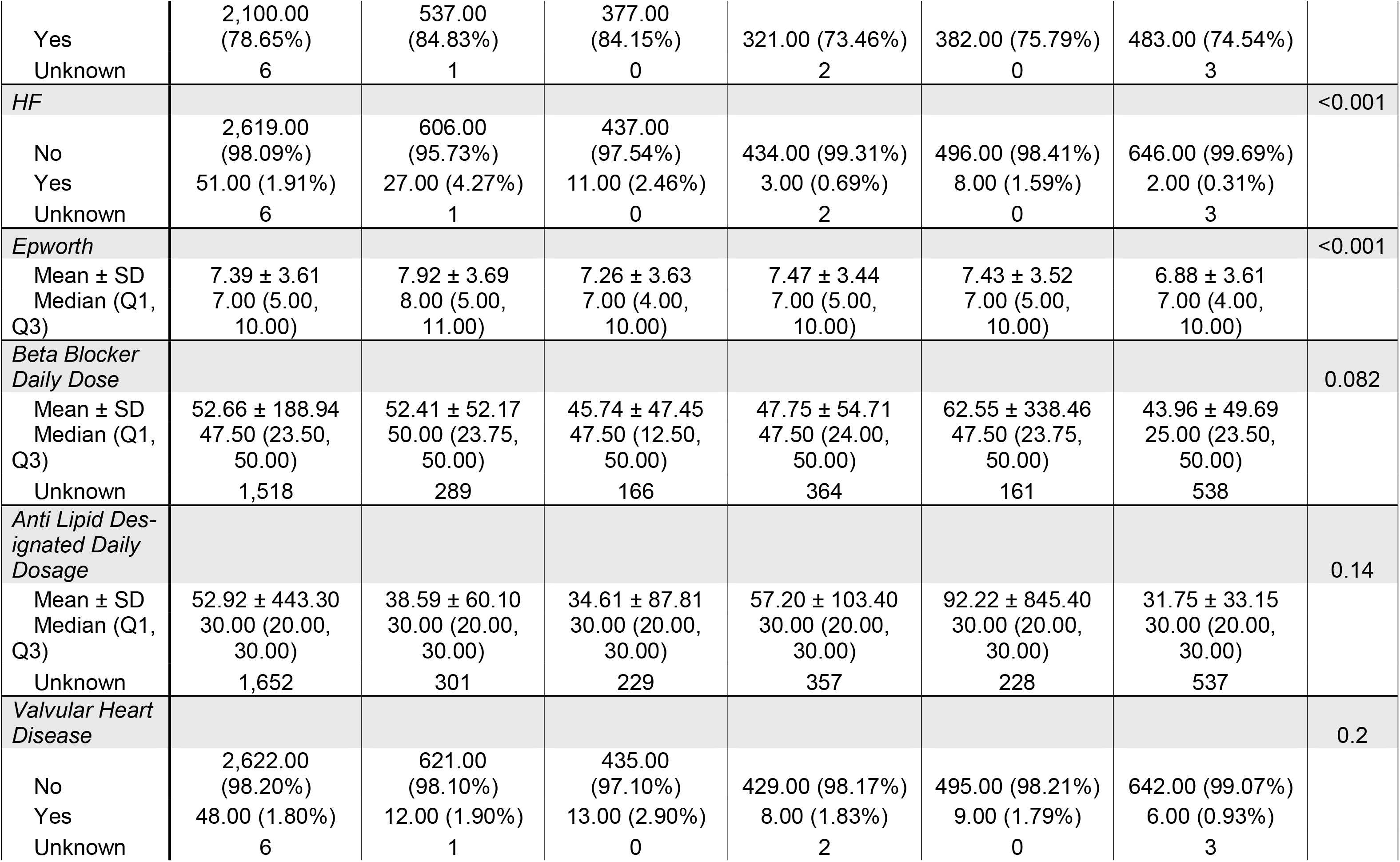
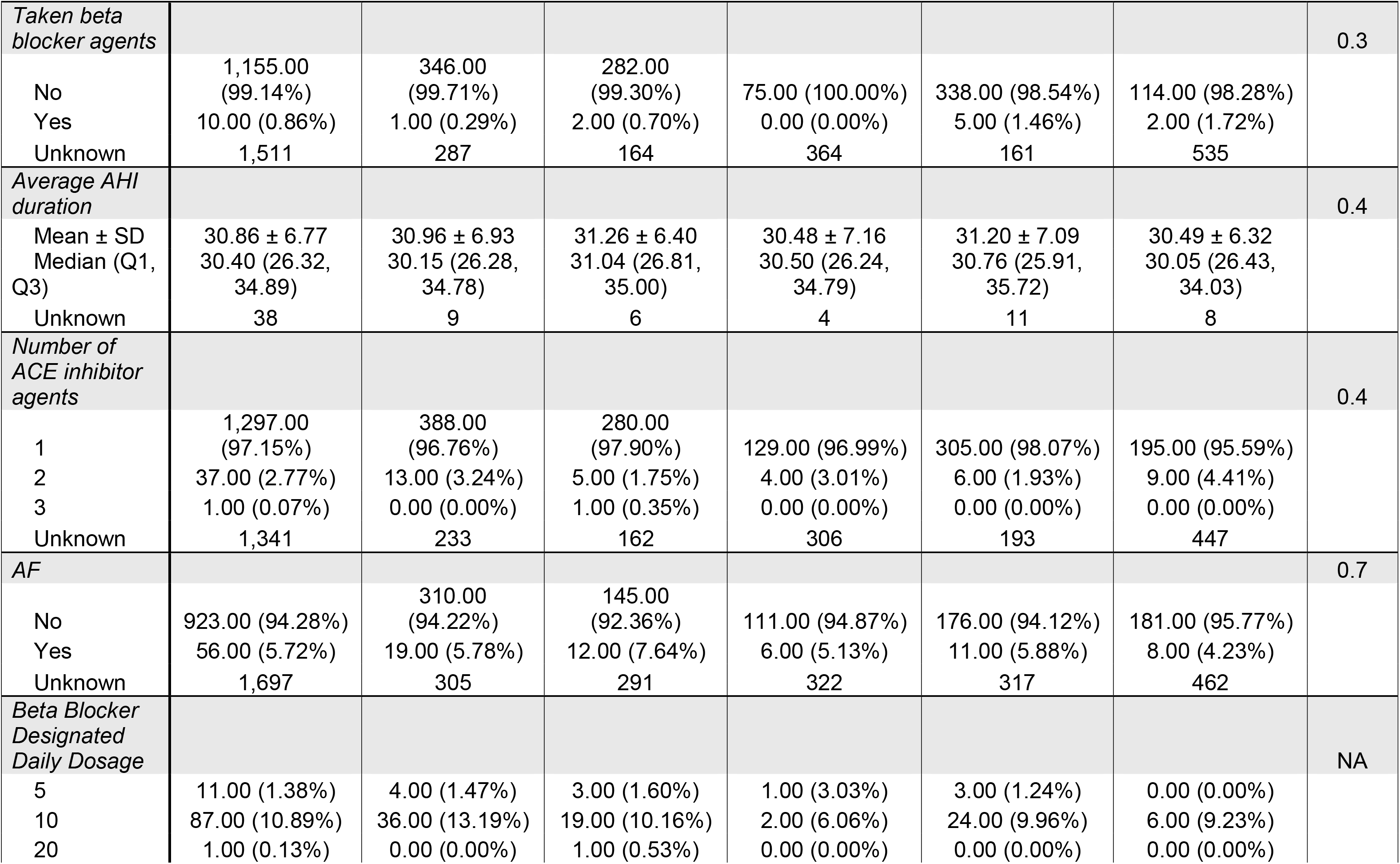
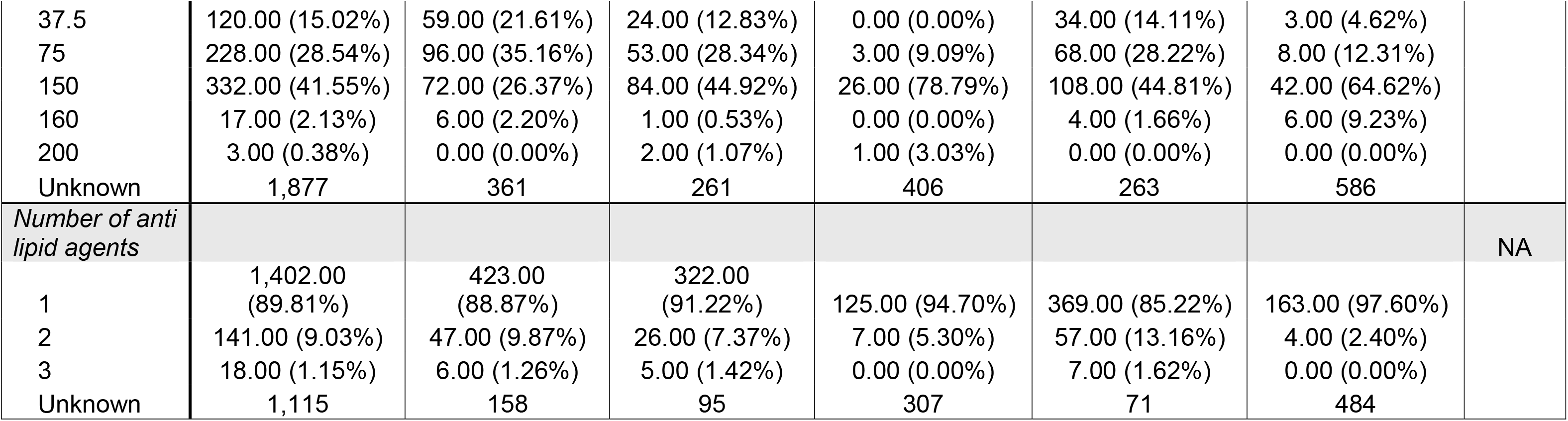
Baseline Characteristics of SAVE Trial Participants Overall and by Obstructive Sleep Apnea Prototype.

### Prototype Development

Using the 10-dimensional UMAP representation, Gaussian mixture models were fitted to evaluate two-through ten-cluster solutions. The C-index supported a five-prototype solution, which is visualized in two dimensions in Figure 2. Boruta random forest–based feature selection was then used to compare each observed variable with randomized shadow variables across 1,000 iterations. Sixty-five variables were identified as statistically significant predictors of prototype membership (p<0.05) (Figure 3A). The top three predictors were diabetes mellitus, prior percutaneous coronary revascularization, and history of consuming >100 cigarettes over the lifetime. Domain-level importance was then derived by aggregating variable-level importance scores within each clinical domain. This analysis showed that substance use, comorbidities, and cardiovascular medication use contributed most strongly to prototype differentiation (Figure 3B).

**Figure 1.**
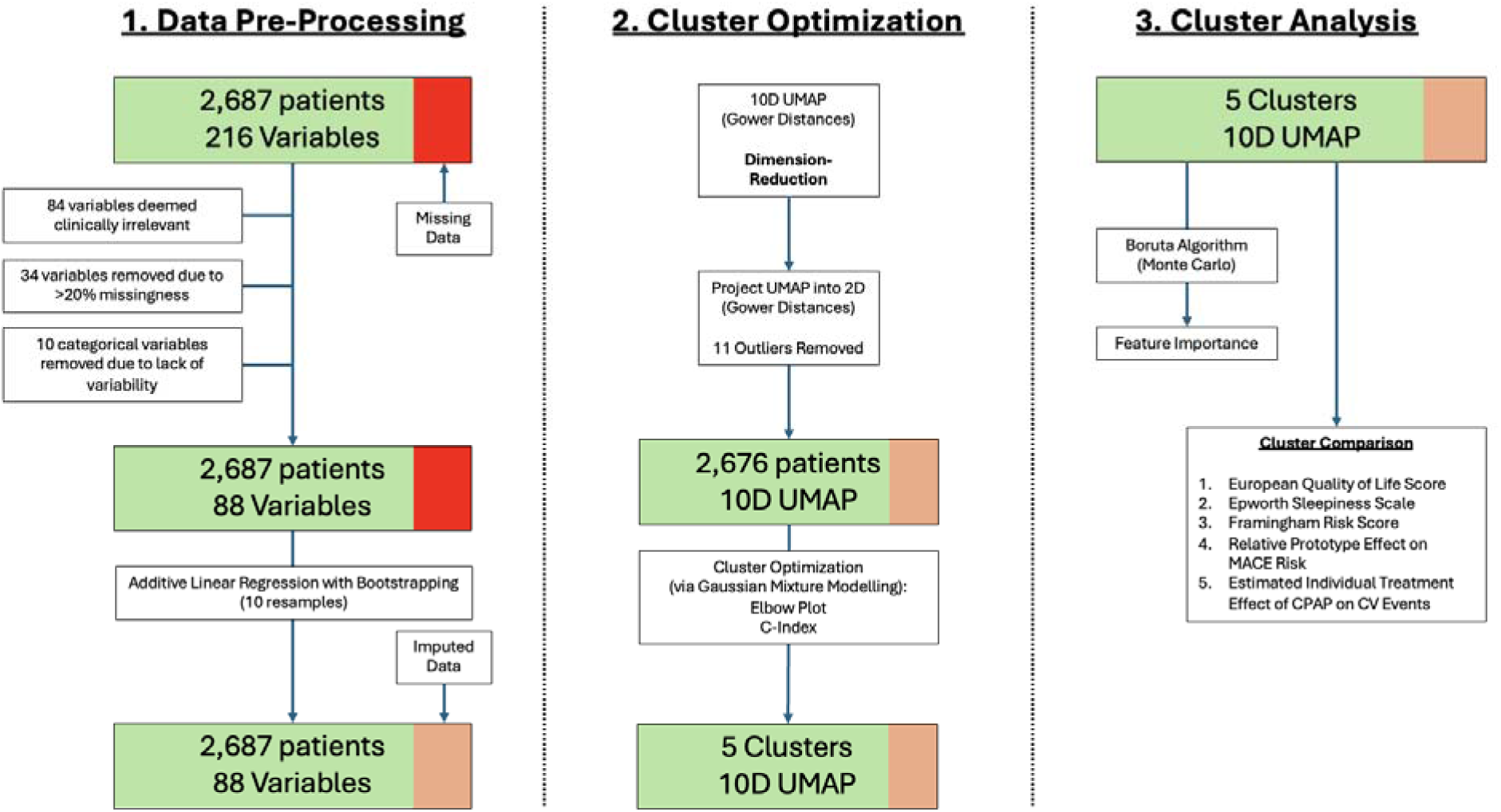
Methods Process Map for Model Building. *Left:* Data Pre-Processing including variable removal and data imputation. *Center:* Cluster Optimization via Uniform Manifold Approximation and Projection (UMAP) and C-Index. *Right:* Cluster Analysis via Boruta Algorithm and Student’s T-Test.

**Figure 2:**
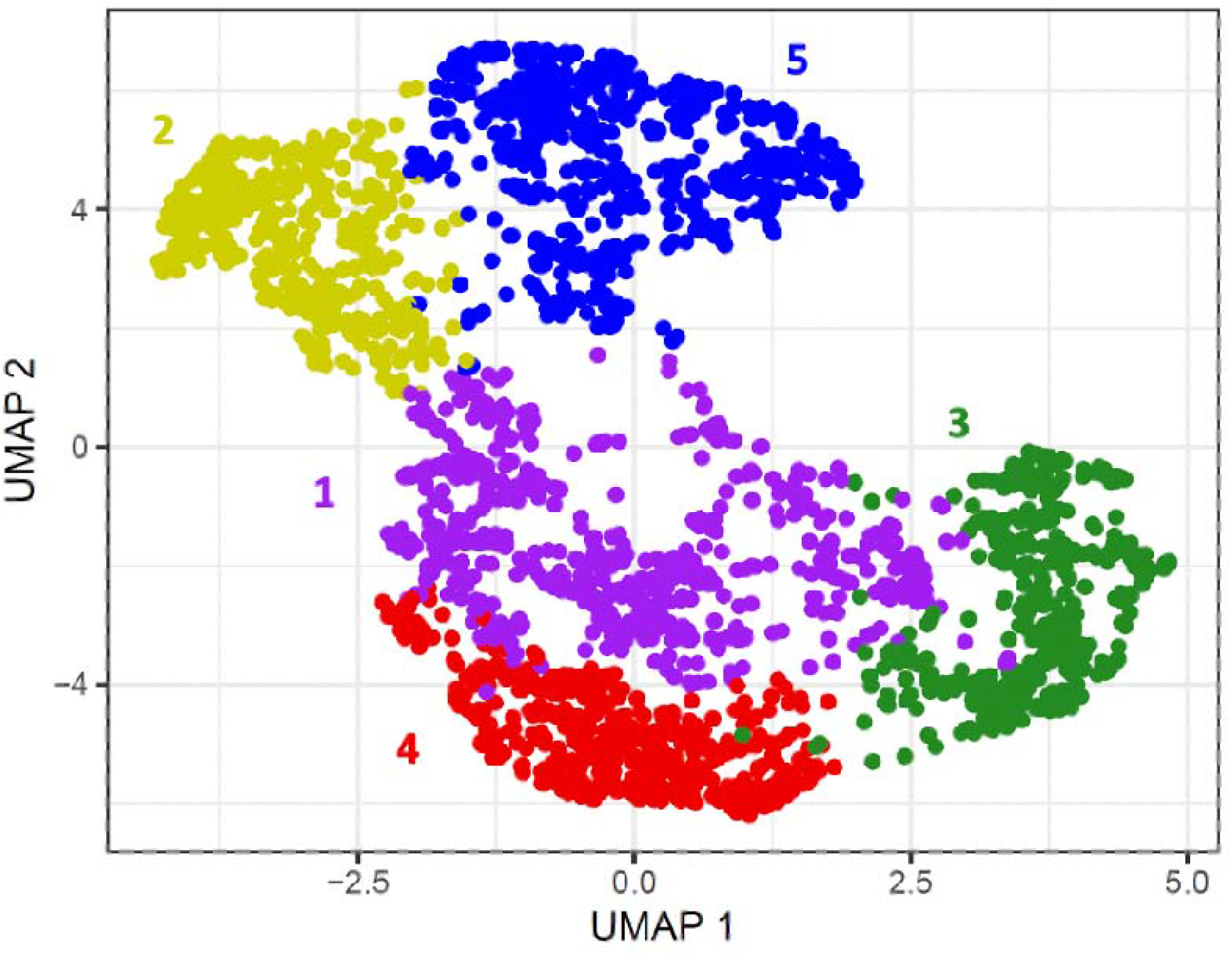
Prototype Membership. Prototypes were developed using a Gaussian mixture model algorithm and optimized to a five-cluster solution. Uniform manifold approximation and projection (UMAP) embeddings, derived from 88 clinically relevant variables, visualize the prototypes in two dimensions, with subjects color-coded by prototype membership.

**Figure 3:**
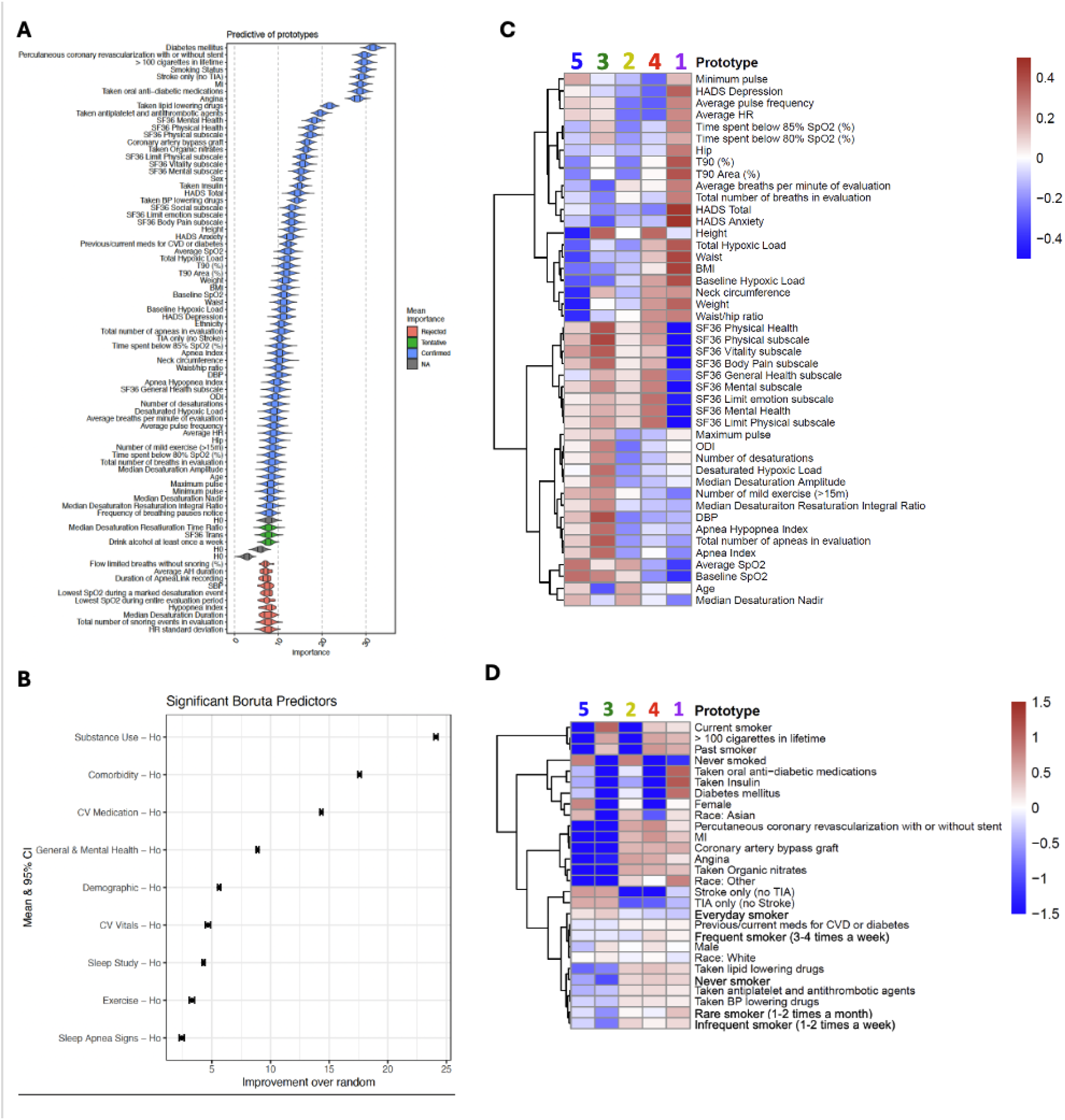
Prototype Feature and Domain Importances. (A) Domain-level importance to identify overarching categories, such as demographic, clinical, and physiological domains, contributing to prototype distinction. (B) We identified variable importance driving prototype classification using additive regression, bootstrapping, and random forest importance scores. (C, D) Heatmaps display the distribution of standardized variable within each prototype. (C) represented by the mean of the continuous variables, with red indicating higher values, while (D) focuses on categorical variables, with red indicating higher proportions. These visualizations underscore key factors driving prototype differentiation.

### Prototype Baseline Characteristics

The final analytic cohort was distributed across the five prototypes as follows: Prototype 1, n=634; Prototype 2, n=448; Prototype 3, n=439; Prototype 4, n=504; and Prototype 5, n=651. Overall differences across prototypes were observed for diabetes mellitus, oral antidiabetic medication use, smoking status, prior myocardial infarction, prior stroke, antiplatelet/antithrombotic therapy, lipid-lowering therapy, sex, BMI, average oxygen saturation, T90, and AHI (all P<0.001) (Table 1). In contrast, valvular heart disease, atrial fibrillation, beta-blocker use, and average apnea-hypopnea duration did not differ across prototypes (all P>0.05).

Across the retained baseline variables, five distinct, clinically interpretable profiles emerged: a metabolic profile in Prototype 1; a never-smoking ischemic profile in Prototype 2; smoking-associated cerebrovascular and coronary profiles in Prototypes 3 and 4, respectively; and a female-and stroke-enriched profile with lower metabolic and coronary comorbidity in Prototype 5 (Figures 3C and 3D; Table 2). Prototype 1 had a high prevalence of diabetes mellitus (483 [76.3%]) and oral antidiabetic medication use (388 [61.2%]). Prototype 2 comprised exclusively never-smokers and had a high prevalence of angina (299 [66.7%]), prior myocardial infarction (234 [52.2%]), and antiplatelet/antithrombotic therapy (415 [92.6%]). Prototypes 3 and 4 consisted almost entirely of participants with current or former smoking histories; Prototype 3 also had a high prevalence of prior stroke (350 [80.1%]), whereas Prototype 4 had the highest prevalence of prior myocardial infarction (333 [66.1%]), antiplatelet/antithrombotic therapy (479 [95.0%]), and lipid-lowering therapy (433 [85.9%]). Prototype 5 had the highest proportion of women (269 [41.3%]) and prior stroke (537 [82.9%]) and the lowest prevalence of prior myocardial infarction (11 [1.7%]) and any heart disease (73 [11.3%]); diabetes mellitus was present in 145 participants (22.4%).

**Table 2.** Summary of Data-Driven Features, MACE Risk, Estimated CPAP Treatment Effects, and Hypothesized Mechanisms Across Obstructive Sleep Apnea Prototypes.

| PROTOTYPE | TOP SAVE TRIAL DATA-DRIVEN PREDICTORS | Baseline Day-time Sleepi-ness | Relative MACE Risk (vs. Overall SAVE Cohort) | Mean Estimated Individual Treat-ment Effect of CPAP on MACE Risk | POSTULATED BIOLOGICAL MECHANISM |
| --- | --- | --- | --- | --- | --- |
| **1 (Purple)** | Diabetes, Metabolism-related medications, Other cardiac meds, Organic nitrates, Insulin | Most sleepy | Highest CV risk | Indication of no Benefit / Possible Harm | CPAP may not address the un-derlying metabolic and in-flammatory drivers of CV dis-ease. |
| **2 (Yellow)** | Smoking-Status Pattern (Never Smoker), Desaturation load, His-tory of stroke, Apnea-hypopnea index, Snoring loudness | Average sleepi-ness | Neutral to high CV risk | No Benefit or Harm | CPAP mitigates hypoxic burden slightly, but lifestyle factors (e.g., smoking) limit protective effect. |
| **3 (Green)** | Blood pressure-lowering drugs, Antiplatelet agents, Angina, Percutaneous coronary revascu-larization, Hypertension | Average sleepi-ness | Neutral to low CV risk | Indication of no Benefit / Possible Harm | CPAP eliminates potential CV benefits of ischemic precondi-tioning among patients treat-ed for coronary artery disease. |
| **4 (Red)** | Smoking history, Antiplatelet agents, Percutaneous coronary revascularization, Angina, Or-ganic nitrates | Average sleepi-ness | Neutral to low CV risk | Indication of no Benefit / Possible Harm | CPAP eliminates potential CV benefits of ischemic precondi-tioning among patients treat-ed for coronary artery disease. |
| **5 (Blue)** | History of stroke, Average ap-nea-hypopnea duration | Least sleepy | Lowest CV risk | Indication of Benefit | CPAP mitigates effects of in-termittent hypoxia on vulnera-ble cerebrovasculature. |

### Symptom Profiles, Cardiovascular Risk, and Predicted CPAP Response

Baseline symptom burden and cardiovascular risk differed across prototypes (Figure 4A–C; Table S2). Prototype 1 had the lowest EQoL utility score (mean, 0.72 [95% CI, 0.69–0.75]), highest baseline ESS (7.92 [7.63–8.21]), and highest baseline FRS (23.3 [22.8–23.9]); each differed significantly from the corresponding values in the other prototypes (all pairwise P<0.05). Prototype 5 had the lowest baseline ESS score (6.88 [6.60–7.16]) and baseline FRS (17.1 [16.5–17.8]; all pairwise P<0.05). Baseline EQoL scores were highest and did not differ between Prototypes 3 and 5, whereas Prototypes 2 and 4 had intermediate scores. Baseline ESS scores were similar across Prototypes 2, 3, and 4, while Prototype 2 had the second-highest baseline FRS (19.8 [19.1–20.5]).

**Figure 4:**
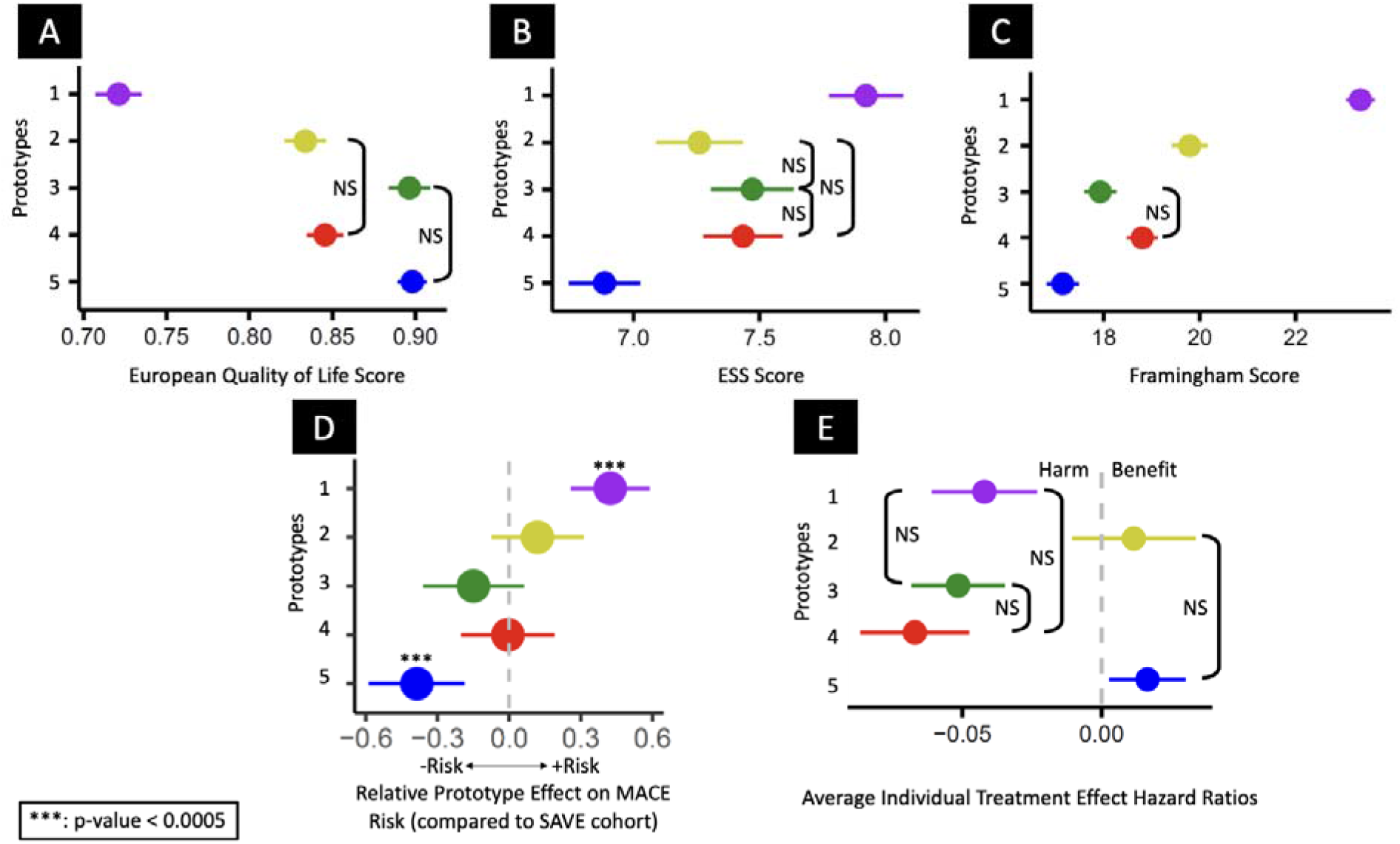
OSA Prototypes by Baseline Symptoms and by MACE Outcomes with CPAP: Mean and standard error of (**A**) health-related quality of life, (**B**) Epworth Sleepiness Score (ESS), and (**C**) Framingham risk score by prototype. (Unless otherwise indicated (labeled NS), pairwise comparisons are significant.) (**D**) Relative Prototype Effect on MACE Risk (compared to SAVE cohort) and (**E**) Estimated individual treatment effects (ITE) of CPAP on CV events across the prototypes, with positive values indicating reduced CV risk and negative values suggesting potential harm.

For the primary composite cardiovascular endpoint, Prototype 1 had the highest relative MACE risk compared with the overall SAVE cohort (0.42 [95% CI, 0.26–0.59]; P<0.0001), whereas Prototype 5 had the lowest (−0.37 [−0.59 to −0.19]; P=0.0002) (Figure 4D). Relative cardiovascular risk did not differ significantly from the overall cohort for Prototypes 2, 3, or 4.

Mean ITE estimates also differed across prototypes (Figure 4E; Table S2). Prototype 5 had a positive mean ITE, consistent with predicted cardiovascular benefit from CPAP (0.02 [95% CI, 0.003– 0.03]). In contrast, mean ITE estimates were negative for Prototype 1 (−0.04 [−0.06 to −0.02]), Prototype 3 (−0.05 [−0.07 to −0.03]), and Prototype 4 (−0.07 [−0.09 to −0.05]), indicating potential predicted harm. Prototype 2 had no statistically significant predicted CPAP effect (0.01 [−0.01 to 0.03]).

## Discussion

Our secondary analysis of the SAVE trial used unsupervised, multidomain machine-learning clustering to identify five distinct OSA prototypes. The prototypes differed across clinical domains and were associated with differences in patient-reported outcomes, cardiovascular risk, and differential treatment responses to CPAP. Baseline EQoL was highest in Prototypes 3 and 5, lowest in Prototype 1, and intermediate in Prototypes 2 and 4. Prototype 1 also had the highest FRS and greatest relative risk of the composite CVD endpoint, whereas Prototype 5 had the lowest values. Mean ITE estimates differed across prototypes, indicating heterogeneity in treatment effect across prototypes. Prototype 5 had a positive mean ITE consistent with predicted cardiovascular benefit from CPAP, whereas Prototypes 1, 3, and 4 had negative mean ITEs consistent with predicted potential harm; Prototype 2 had no statistically significant predicted treatment effect. To our knowledge, these findings are the first to demonstrate that comprehensive, multidimensional clustering across substance use, comorbidities, cardiovascular medication use, general and mental health, demographics, cardiovascular vital signs, sleep-study measures, exercise, and sleep apnea-related symptoms can identify distinct OSA prototypes with differential treatment response to CPAP.

Although heterogeneity within OSA has been explored across individual domains using prespecified hypotheses, this analysis demonstrates how information across nine routinely available baseline clinical domains can be integrated into more complete, data-driven patient profiles. These multidimensional differences yielded clinically interpretable profiles with distinct baseline cardiovascular risk, and the impact of CPAP therapy on cardiovascular events was not uniform but instead varied by prototype. Taken together, these findings suggest that a “one-size-fits-all” approach to OSA management may be suboptimal. Following external validation, prototype-informed approaches could guide prospective trial enrichment and, ultimately, help identify patients most likely to derive cardiovascular benefit from CPAP that may be obscured when all patients with an elevated AHI are treated as a single population.

Our findings align with and may help reconcile prior evidence from randomized trials and observational studies of OSA and cardiovascular risk. Like SAVE, other secondary prevention trials in predominantly non-sleepy OSA populations, including RICCADSA in stable coronary artery disease and ISAACC in acute coronary syndrome, found no overall reduction in cardiovascular events with CPAP therapy (6–8). These neutral findings contrast with observational cohort studies in which untreated severe OSA was associated with higher cardiovascular event rates and CPAP use was associated with lower cardiovascular risk (Marín et al., 2005; Campos-Rodríguez et al., 2012). Differences in trial populations may partly explain these findings. In addition to excluding profoundly sleepy patients for ethical reasons, these RCTs generally enrolled patients with OSA defined by an AHI4% >15 events/hour. Given the substantial heterogeneity within OSA, broad enrollment criteria may have diluted potential treatment benefit among subgroups more likely to respond to CPAP. The prototypes developed in this study define multidimensional patient profiles that could support a precision approach to trial enrollment to test the effects of OSA treatment within specific subgroups.

This study extends the growing literature on OSA phenotyping by examining the relationship between multidomain patient profiles, cardiovascular risk, and heterogeneous treatment response to CPAP. Prior studies have identified OSA subgroups using narrower symptom, clinical, sleep-study, or physiologic feature sets (12, 18, 19). Latent class and clustering analyses have described phenotypes ranging from minimally symptomatic to highly symptomatic OSA and polysomnographic profiles associated with differential cardiovascular risk (9, 11). Biomarker and imaging-based clustering has also identified inflammatory OSA subtypes with differential changes in protein expression after CPAP therapy (20, 21). However, few studies have integrated a comparably broad set of routinely available clinical and sleep-study variables to comprehensively define OSA prototypes, and to our knowledge, none have examined heterogeneity of cardiovascular treatment effects of CPAP across subgroups using causal inference methods. In a post hoc machine-learning analysis of the ISAACC trial, model-derived subgroups had heterogeneous estimated CPAP treatment effects on cardiovascular outcomes, with findings consistent with benefit in some subgroups and potential harm in others (22). Together, these studies support further evaluation of multidomain phenotyping as an approach to link OSA profiles with symptoms, quality of life, cardiovascular risk, and potential variation in CPAP responsiveness. Building on this work, our study used unsupervised clustering of nine routinely available baseline clinical domains in a randomized trial cohort to identify OSA prototypes associated with distinct symptom burden, quality of life, cardiovascular risk, and heterogeneous treatment responses to CPAP.

Several limitations should be considered. First, the clustering and subgroup findings were derived post hoc from a single trial cohort. As with other data-driven phenotyping approaches, the identified prototypes depend on the specific variables and analytic methods used and require replication in independent cohorts before broader application. In addition, prototype-specific mean ITE estimates are model-based, counterfactual predictions rather than directly observed treatment effects and require prospective validation. Second, mean CPAP adherence in SAVE was modest (3.3 ± 2.3 hours/night), which may have attenuated treatment effects in the randomized trial. Third, the SAVE population comprised predominantly older men with established cardiovascular disease and without excessive daytime sleepiness, which may limit generalizability to typical sleep clinic populations. Accordingly, these findings may not apply to primary prevention settings, younger or more symptomatic patients, or populations with different demographic characteristics. Finally, prototype derivation was limited to routinely available clinical and sleep-study variables; other factors, including genetic susceptibility and specific pathophysiologic traits, may also influence treatment responsiveness but are not captured within the available data. These limitations temper interpretation of our findings and underscore the need for prospective studies to replicate the prototypes and determine whether model-predicted treatment heterogeneity corresponds to observed CPAP effects.

## Conclusion

Using unsupervised clustering of 88 routinely available baseline variables spanning nine clinical domains within the SAVE randomized trial cohort, we identified five OSA prototypes that differed in symptom burden, quality of life, cardiovascular risk, and heterogeneous predicted responsiveness to CPAP. This comprehensive multidomain framework moves beyond isolated sleep metrics and average treatment effects to capture clinically relevant complexity within OSA. Although CPAP did not reduce cardiovascular events in the overall SAVE cohort, the prototype-specific ITE patterns suggest that important variation in treatment benefit may be obscured when OSA is treated as a single population. Following replication in independent cohorts and prospective validation, these prototypes could provide a pragmatic foundation for enriching future trials and advancing precision approaches to identify patients most likely to derive cardiovascular benefit from CPAP.

## Author Contributions

**Tahir Hafeez Malik:** Conceptualization, Data curation, Formal analysis, Methodology, Writing – original draft, Writing – review & editing

**David Daniel:** Writing – original draft

**Jo Hsuan Lee:** Data curation, Formal analysis, Methodology

**Zainab Al-Taie:** Data curation, Formal analysis, Methodology

**Kavya Devarakonda:** Writing – original draft

**Samira Khan:** Conceptualization, Data curation

**Vi Le:** Conceptualization, Data curation

**Kelly A. Loffler:** Conceptualization, Data curation, Writing – original draft

**Craig S. Anderson:** Conceptualization, Data curation, Writing – original draft

**Mathias Baumert:** Conceptualization, Data curation, Writing – original draft

**Jason C. Kovacic:** Conceptualization, Data curation, Writing – original draft

**Shishir Adhikari:** Conceptualization, Data curation, Writing – original draft

**Vaishnavi Kundel:** Conceptualization, Data curation, Formal analysis, Methodology, Writing – original draft, Writing – review & editing

**Cameron S. McAlpine:** Conceptualization, Data curation, Writing – original draft

**Girish Nadkarni:** Conceptualization, Data curation, Formal analysis, Methodology, Writing – original draft

**Oren Cohen:** Conceptualization, Data curation, Formal analysis, Methodology, Writing – original draft, Writing – review & editing

**Mayte Suárez-Fariñas:** Conceptualization, Data curation, Formal analysis, Methodology, Writing – original draft, Writing – review & editing

**Neomi Shah:** Conceptualization, Data curation, Formal analysis, Methodology, Writing – original draft, Writing – review & editing

## Conflicts of Interest

T.H.M., D.D., J.H.L., Z.A., K.D., S.K., V.L., K.A.L., C.S.A., M.B., J.C.K., S.A., V.K., C.S.M., G.N., O.C., M.S., and N.S. reported no relevant financial relationships with ineligible companies.

## Data Availability

All data produced in the present study are available upon reasonable request to the authors.

## Notes

### Competing Interest Statement

The authors have declared no competing interest.

### Clinical Trial

NCT00738179

## References

1. Yeghiazarians Y, Jneid H, Tietjens JR, Redline S, Brown DL, El-Sherif N, et al. Obstructive Sleep Apnea and Cardiovascular Disease: A Scientific Statement from the American Heart Association. Circulation 2021;

2. Campos-Rodriguez F, Martinez-Garcia MA, De La Cruz-Moron I, Almeida-Gonzalez C, Catalan-Serra P, Montserrat JM. Cardiovascular Mortality in Women With Obstructive Sleep Apnea With or Without Continuous Positive Airway Pressure Treatment A Cohort Study. 2012. at <https://annals.org>.

3. Gottlieb DJ, Yenokyan G, Newman AB, O’Connor GT, Punjabi NM, Quan SF, et al. Prospective study of obstructive sleep apnea and incident coronary heart disease and heart failure: The sleep heart health study. Circulation 2010;122:352–360.

4. Marin JM, Carrizo SJ, Vicente E, Agusti AGN. Long-term cardiovascular outcomes in men with obstructive sleep apnoea-hypopnoea with or without treatment with continuous positive airway pressure: an observational study. Lancet 365:1046–53.

5. Benjafield A V, Ayas NT, Eastwood PR, Heinzer R, Ip MSM, Morrell MJ, et al. Estimation of the global prevalence and burden of obstructive sleep apnoea: a literature-based analysis. Lancet Respir Med 2019;7:687–698.

6. McEvoy RD, Antic NA, Heeley E, Luo Y, Ou Q, Zhang X, et al. CPAP for Prevention of Cardiovascular Events in Obstructive Sleep Apnea. New England Journal of Medicine 2016;375:919–931.

7. Peker Y, Glantz H, Eulenburg C, Wegscheider K, Herlitz J, Thunström E. Effect of positive airway pressure on cardiovascular outcomes in coronary artery disease patients with nonsleepy obstructive sleep apnea: The RICCADSA randomized controlled trial. Am J Respir Crit Care Med 2016;194:613–620.

8. Sánchez-de-la-Torre M, Sánchez-de-la-Torre A, Bertran S, Abad J, Duran-Cantolla J, Cabriada V, et al. Effect of obstructive sleep apnoea and its treatment with continuous positive airway pressure on the prevalence of cardiovascular events in patients with acute coronary syndrome (ISAACC study): a randomised controlled trial. Lancet Respir Med 2020;8:359–367.

9. Mazzotti DR, Keenan BT, Lim DC, Gottlieb DJ, Kim J, Pack AI. Symptom subtypes of obstructive sleep apnea predict incidence of cardiovascular outcomes. Am J Respir Crit Care Med 2019;200:493–506.

10. Ye L, Pien GW, Ratcliffe SJ, Björnsdottir E, Arnardottir ES, Pack AI, et al. The different clinical faces of obstructive sleep apnoea: A cluster analysis. European Respiratory Journal 2014;44:1600–1607.

11. Zinchuk A V., Jeon S, Koo BB, Yan X, Bravata DM, Qin L, et al. Polysomnographic phenotypes and their cardiovascular implications in obstructive sleep apnoea. Thorax 2018;73:472–480.

12. Azarbarzin A, Sands SA, Stone KL, Taranto-Montemurro L, Messineo L, Terrill PI, et al. The hypoxic burden of sleep apnoea predicts cardiovascular disease-related mortality: The Osteoporotic Fractures in Men Study and the Sleep Heart Health Study. Eur Heart J 2019;40:1149–1157a.

13. Azarbarzin A, Sands SA, Younes M, Taranto-Montemurro L, Sofer T, Vena D, et al. The sleep apnea-specific pulse-rate response predicts cardiovascular morbidity and mortality. Am J Respir Crit Care Med 2021;203:1546–1555.

14. Cohen O, Kundel V, Robson P, Al-Taie Z, Suárez-Fariñas M, Shah NA. Achieving Better Understanding of Obstructive Sleep Apnea Treatment Effects on Cardiovascular Disease Outcomes through Machine Learning Approaches: A Narrative Review. J Clin Med 2024;

15. Cohen O, Kundel V, Yaggi HK, Shah NA, Alcantara C, Azarbarzin A, et al. The Great Controversy of Obstructive Sleep Apnea Treatment for Cardiovascular Risk Benefit: Advancing the Science through Expert Consensus An Official American Thoracic Society Workshop Report. Ann Am Thorac Soc 2025;22:1–22.

16. Hubert LJ, Levin JR. A general statistical framework for assessing categorical clustering in free recall. Psychol Bull 1976;83:1072–1080.

17. Cohen O, Al-Taie Z, Kundel V, Khan S, Devarakonda K, Le V, et al. Individualized treatment effects of CPAP on secondary cardiovascular outcomes in non-sleepy obstructive sleep apnea patients. Communications Medicine 2026;doi:10.1038/s43856-026-01457-1.

18. Pinilla L, Esmaeili N, Labarca G, Martinez-Garcia MÁ, Torres G, Gracia-Lavedan E, et al. Hypoxic burden to guide CPAP treatment allocation in patients with obstructive sleep apnoea: a post hoc study of the ISAACC trial. European Respiratory Journal 2023;62:.

19. Azarbarzin A, Zinchuk A, Wellman A, Labarca G, Vena D, Gell L, et al. Cardiovascular Benefit of Continuous Positive Airway Pressure in Adults with Coronary Artery Disease and Obstructive Sleep Apnea without Excessive Sleepiness. Am J Respir Crit Care Med 2022;206:767–774.

20. Kundel V, Cohen O, Khan S, Patel M, Kim-Schulze S, Kovacic J, et al. Advanced Proteomics and Cluster Analysis for Identifying Novel Obstructive Sleep Apnea Subtypes before and after Continuous Positive Airway Pressure Therapy. Ann Am Thorac Soc 2023;20:1038– 1047.

21. Cohen O, Kaufman AE, Choi H, Khan S, Robson PM, Suárez-Fariñas M, et al. Pharyngeal Inflammation on Positron Emission Tomography/Magnetic Resonance Imaging Before and After Obstructive Sleep Apnea Treatment. Ann Am Thorac Soc 2023;20:574–583.

22. Cohen O, Al-Taie Z, Kundel V, Khan S, Devarakonda K, Robson PM, et al. Individualized Treatment Effects of CPAP in Obstructive Sleep Apnea: A Post-hoc Machine Learning Analysis in Save. A19 THE FRONTLINE SLEEP APNEA: INNOVATIONS IN OSA American Thoracic Society; 2024. p. A1058–A1058.doi:10.1164/ajrccm-conference.2024.209.1_MeetingAbstracts.A1058.

