## Supplementary material for "Prototyping OSA: A New Framework for Predicting Cardiovascular Outcomes and CPAP Benefit": Online Supplement

**Title:** Prototyping OSA: A New Framework for Predicting Cardiovascular Outcomes and CPAP Benefit – Online Supplement

**Figure S1.** SAVE cohort distribution across 2-Dimensional UMAPs (Top, Middle, Bottom) and outlier identification

**Table S1**. All Variables and Reasons for Exclusion

**Table S2.** European Quality of Life Score, Epworth Sleepiness Scale, Framingham Risk Score, and Average Individual Treatment Effect by Prototype

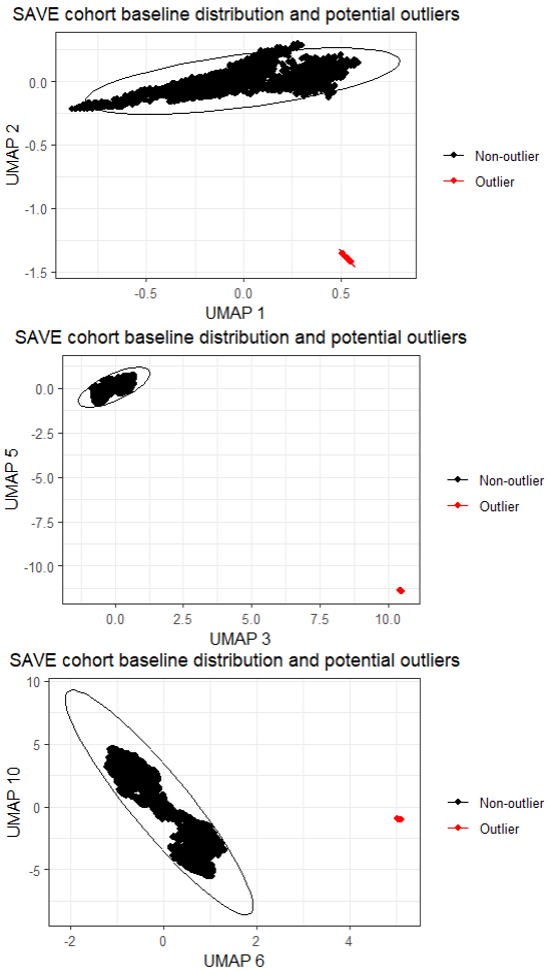

**Figure S1.** SAVE cohort distribution across 2-Dimensional UMAPs (Top, Middle, Bottom) and outlier identification

**Table S1**. All Variables and Reasons for Exclusion

| **Definition** | **Considered for Prototype (1 = Yes,**  **0 = No)** | **Category** | **Used for Prototype (1 = Yes,**  **0 = No)** | **Reason for exclusion** |
| --- | --- | --- | --- | --- |
| Matched Propensity Score from Model 1 | 0 | NA | NA | NA |
| Subject ID | 0 | NA | NA | NA |
| SAVE_ApneaLink_Baseline.csv_SubjectNumberStr | 0 | NA | NA | NA |
| NA | 0 | NA | NA | NA |
| NA | 0 | NA | NA | NA |
| NA | 0 | NA | NA | NA |
| NA | 0 | NA | NA | NA |
| Taken Insulin | 1 | CV Medication | 1 | NA |
| Taken Organic Nitrates | 0 | CV Medication | NA | NA |
| Taken other Cardiac Medications | 1 | CV Medication | 1 | NA |
| Taken traditional Chinese Medicines | 0 | Medication | NA | NA |
| Age | 1 | Demographic | 1 | NA |
| Apnea Index | 1 | Sleep Study | 1 | NA |
| Apnea Hypopnea Index | 1 | Sleep Study | 1 | NA |
| Oxygen Desaturation Index | 0 | NA | NA | NA |
| Total Number of Apneas in Evaluation | 1 | Sleep Study | 1 | NA |
| Average AH Duration | 1 | Sleep Study | 1 | NA |
| Average Breaths per Minute of Evaluation | 1 | Sleep Study | 1 | NA |
| Average Pulse Frequency | 1 | Sleep Study | 1 | NA |
| Average SpO2 | 0 | Sleep Study | NA | NA |
| Average SpO2 | 1 | Sleep Study | 1 | NA |
| Beer Consumption per Week | 0 | Substance Use | NA | NA |
| Body Massi Index | 1 | Demographic | 1 | NA |
| Diastolic Blood Pressure | 1 | CV Vitals | 1 | NA |
| HDL | 1 | Laboratory | 0 | >20% missing |
| Height | 1 | Demographic | 1 | NA |
| Hip | 1 | Demographic | 1 | NA |
| Heart Rate | 0 | CV Vitals | NA | NA |
| LDL | 1 | Laboratory | 0 | >20% missing |
| Neck Circumference | 1 | Demographic | 1 | NA |
| Number of Antihypertensive Drug Agents | 1 | CV Medication | 0 | >20% missing |
| Number of Diuretics Agents | 1 | CV Medication | 0 | >20% missing |
| Number of Beta Blocking Agents | 1 | CV Medication | 0 | >20% missing |
| Number of Calcium Channel Blockers Agents | 1 | CV Medication | 0 | >20% missing |
| Number of ACE Inhibitor Agents | 1 | CV Medication | 0 | >20% missing |
| Number of Lipid Modifying Agents | 1 | CV Medication | 0 | >20% missing |
| Number of Blood Glucose Lowering Drug Agents | 1 | CV Medication | 0 | >20% missing |
| Systolic Blood Pressure | 1 | CV Vitals | 1 | NA |
| Spirit Consumption per Week | 0 | Substance Use | NA | NA |
| Sum of Antihypertensive Drugs Daily Dose | 1 | CV Medication | 0 | >20% missing |
| Sum of Diuretics Daily Dose | 1 | CV Medication | 0 | >20% missing |
| Sum of Beta Blocking Agents Daily Dose | 1 | CV Medication | 0 | >20% missing |
| Sum of Calcium Channel Blockers Daily Dose | 1 | CV Medication | 0 | >20% missing |
| Sum of ACE Inhibitors Daily Dose | 1 | CV Medication | 0 | >20% missing |
| Sum of Lipid Modifying Agents Daily Dose | 1 | CV Medication | 0 | >20% missing |
| Sum of Blood Glucose Lowering Drugs Daily Dose | 1 | CV Medication | 0 | >20% missing |
| Sum of Antihypertensive Drugs Designated Daily Dosage | 1 | CV Medication | 0 | >20% missing |
| Sum of Diuretics Designated Daily Dosage | 1 | CV Medication | 0 | >20% missing |
| Sum of Beta Blocking Agents Designated Daily Dosage | 1 | CV Medication | 0 | >20% missing |
| Sum of Calcium Channel Blockers Designated Daily Dosage | 1 | CV Medication | 0 | >20% missing |
| Sum of ACE Inhibitors Designated Daily Dosage | 1 | CV Medication | 0 | >20% missing |
| Sum of Lipid Modifying Agents Designated Daily Dosage | 1 | CV Medication | 0 | >20% missing |
| Sum of Blood Glucose Lowering Drugs Designated Daily Dosage | 1 | CV Medication | 0 | >20% missing |
| Antihypertensive Drugs Ratio Dose/Designated Daily Dosage | 1 | CV Medication | 0 | >20% missing |
| Diuretics: Ratio Dose/Designated Daily Dosage at Baseline | 0 | CV Medication | NA | NA |
| Beta Blocking Agents: Ratio Dose/Designated Daily Dosage at Baseline | 0 | CV Medication | NA | NA |
| Calcium Channel Blockers: Ratio dose/Designated Daily Dosage at Baseline | 0 | CV Medication | NA | NA |
| ACE Inhibitors: Ratio Dose/Designated Daily Dosage at Baseline | 0 | CV Medication | NA | NA |
| Lipid Modifying Agents: Ratio Dose/Designated Daily Dosage at Baseline | 0 | CV Medication | NA | NA |
| Blood Glucose Lowering Drugs: Ratio Dose/Designated Daily Dosage at Baseline | 0 | CV Medication | NA | NA |
| Total Cholesterol | 1 | Laboratory | 0 | >20% missing |
| Triglycerides | 1 | Laboratory | 0 | >20% missing |
| Waist | 1 | Demographic | 1 | NA |
| Waist/Hip Ratio | 1 | Demographic | 1 | NA |
| Weight | 1 | Demographic | 1 | NA |
| Wine Consumption per Week | 0 | Substance Use | NA | NA |
| Baseline Hypoxic Load | 1 | Sleep Study | 1 | NA |
| Baseline SpO2 | 0 | Sleep Study | NA | NA |
| Baseline SpO2 | 1 | Sleep Study | 1 | NA |
| Taken BP Lowering Drugs | 1 | CV Medication | 1 | NA |
| Taken Lipid Lowering Drugs | 1 | CV Medication | 1 | NA |
| Taken Oral Anti-Diabetic Medications | 1 | CV Medication | 1 | NA |
| Taken Antiplatelet and Antithrombotic Agents | 1 | CV Medication | 1 | NA |
| Taken Alimentary Tract and Metabolism Medications | 1 | CV Medication | 1 | NA |
| Total Number of Breaths in Evaluation | 1 | Sleep Study | 1 | NA |
| Cheyne Stokes Epochs (%) | 1 | Sleep Study | 1 | NA |
| Total Cheyne Stokes Epochs | 0 | Sleep Study | NA | NA |
| Desaturated Hypoxic Load | 1 | Sleep Study | 1 | NA |
| Duration of Flow Analysis | 0 | Sleep Study | NA | NA |
| Duration of ApneaLink Recording | 1 | Sleep Study | 1 | NA |
| European Quality of Life Utility Score | 0 | Sleep Symptom | NA | NA |
| Epworth Sleepiness Score | 0 | Sleep Symptom | NA | NA |
| CAD Only | 0 | Comorbidity | NA | NA |
| TIA or Stroke | 0 | Comorbidity | NA | NA |
| Flag of TIA/STROKE/CAD; Medical History | 0 | Comorbidity | NA | NA |
| Flag of TIA only; Medical History | 0 | Comorbidity | NA | NA |
| Total Flow Limited Breaths without Snoring | 0 | Sleep Study | NA | NA |
| Flow Limited Breaths without Snoring (%) | 1 | Sleep Study | 1 | NA |
| Total Flow Limited Breaths with Snoring | 0 | Sleep Study | NA | NA |
| Flow Limited Breaths with Snoring (%) | 1 | Sleep Study | 1 | NA |
| Hospital Anxiety and Depression Score: Anxiety | 1 | General & Mental Health | 1 | NA |
| Hospital Anxiety and Depression Score: Depression | 1 | General & Mental Health | 1 | NA |
| Hospital Anxiety and Depression Score: Total | 1 | General & Mental Health | 1 | NA |
| HR IQR | 0 | Sleep Study | NA | NA |
| Average HR | 1 | Sleep Study | 1 | NA |
| Median HR | 0 | Sleep Study | NA | NA |
| SAVE_MatlabCustom.csv_HRQ2bpm | 0 | Sleep Study | NA | NA |
| 75th Percentile HR | 0 | Sleep Study | NA | NA |
| HR Standard Deviation | 1 | Sleep Study | 1 | NA |
| Hypopnea Index | 1 | Sleep Study | 1 | NA |
| Total Number of Hypopneas in Evaluation | 0 | Sleep Study | NA | NA |
| Oxygen Desaturation Index | 0 | Sleep Study | NA | NA |
| Apnea-Hypopnea Index | 0 | Sleep Study | NA | NA |
| Calculated Epworth Sleepiness Score at screening | 0 | Symptoms | NA | NA |
| Treatment as Randomised | 0 | NA | NA | NA |
| Allocated Treatment Code | 0 | NA | NA | NA |
| Drink Alcohol at Least once a Week | 1 | Substance Use | 1 | NA |
| Average Beer per Week | 1 | Substance Use | 0 | >20% missing |
| Average Spirits per Week | 1 | Substance Use | 0 | >20% missing |
| Average Wine per Week | 1 | Substance Use | 0 | >20% missing |
| Number of Mild Exercise (>15m) | 1 | Exercise | 1 | NA |
| Number of Moderate Exercise (>15m) | 1 | Exercise | 1 | NA |
| Number of Vigorous Exercise (>15m) | 1 | Exercise | 1 | NA |
| Number of Cigarettes/day | 1 | Substance Use | 0 | >20% missing |
| Smoke Cigarettes Every Day | 1 | Substance Use | 0 | >20% missing |
| Smoke Cigars Every Day | 1 | Substance Use | 0 | Rare or Invariant |
| Number of Cigars/Day | 1 | Substance Use | 0 | >20% missing |
| Smoke Cigarettes after Awake for 30 minutes | 1 | Substance Use | 0 | >20% missing |
| > 100 Cigarettes in Lifetime | 1 | Substance Use | 1 | NA |
| Time since Last Cigarette | 1 | Substance Use | 0 | Rare or Invariant |
| Pipe/Day | 1 | Substance Use | 0 | >20% missing |
| Current Pipe Smoker | 1 | Substance Use | 0 | Rare or Invariant |
| Established Coronary Disease confirmed as Eligibility Criterion | 0 | Comorbidity | NA | NA |
| Established Cerebrovascular Disease confirmed as Eligibility Criterion | 0 | Comorbidity | NA | NA |
| Atrial Fibrillation | 1 | Comorbidity | 0 | Rare or Invariant |
| Angina | 1 | Comorbidity | 1 | NA |
| Hypertension | 1 | Comorbidity | 1 | NA |
| Coronary Artery Bypass Graft | 1 | Comorbidity | 1 | NA |
| Carotid Endarterectomy or Percutaneous Stent | 1 | Comorbidity | 0 | Rare or Invariant |
| Previous or Current Medication for Cardiovascular Disease or Diabetes | 1 | Comorbidity | 1 | NA |
| Other form of Cardiovascular Disease | 0 | Comorbidity | NA | NA |
| Diabetes Mellitus | 0 | Comorbidity | NA | NA |
| Any Heart Disease | 0 | Comorbidity | NA | NA |
| Heart Failure | 1 | Comorbidity | 0 | Rare or Invariant |
| Intracerebral Stent | 0 | Comorbidity | NA | NA |
| Myocardial Infarction | 1 | Comorbidity | 1 | NA |
| Percutaneous Coronary Revascularization With or Without Stent | 1 | Comorbidity | 1 | NA |
| Stroke | 0 | Comorbidity | NA | NA |
| TIA | 0 | Comorbidity | NA | NA |
| Valvular Heart Disease | 1 | Comorbidity | 0 | Rare or Invariant |
| Ethnicity | 0 | NA | NA | NA |
| Ethnicity | 1 | Demographic | 1 | NA |
| Sex | 1 | Demographic | 1 | NA |
| Snoring Bothers Others | 1 | Sleep Apnea Signs | 1 | NA |
| Snoring Frequency | 1 | Sleep Apnea Signs | 0 | Rare or Invariant |
| Snoring Loudness | 1 | Sleep Apnea Signs | 1 | NA |
| Snorer | 1 | Sleep Apnea Signs | 0 | Rare or Invariant |
| Frequency of Breathing Pauses Notice | 1 | Sleep Apnea Signs | 1 | NA |
| Lowest SpO2 during a Marked Desaturation Event | 1 | Sleep Study | 1 | NA |
| Lowest SpO2 during Entire Evaluation Period | 1 | Sleep Study | 1 | NA |
| Maximum Pulse | 1 | Sleep Study | 1 | NA |
| SF36 Mental Health | 1 | General & Mental Health | 1 | NA |
| SAVE_MatlabCustom.csv_meanNNms | 0 | Sleep Study | NA | NA |
| SAVE_EventDurations.csv_medAHDuration | 0 | Sleep Study | NA | NA |
| Median Desaturation Amplitude | 1 | Sleep Study | 1 | NA |
| Median Desaturation Duration | 1 | Sleep Study | 1 | NA |
| Median Desaturation Integral | 0 | Sleep Study | NA | NA |
| Median Desaturation Nadir | 1 | Sleep Study | 1 | NA |
| Median Desaturaiton Resaturation Integral Ratio | 1 | Sleep Study | 1 | NA |
| Median Desaturation Resatiuration Time Ratio | 1 | Sleep Study | 1 | NA |
| Minimum Pulse | 1 | Sleep Study | 1 | NA |
| Number of Desaturations | 1 | Sleep Study | 1 | NA |
| Number of Desaturations | 0 | Sleep Study | NA | NA |
| ODI | 1 | Sleep Study | 1 | NA |
| SF36 Physical Health | 1 | General & Mental Health | 1 | NA |
| NA | 0 | NA | NA | NA |
| SAVE_MatlabCustom.csv_rmssdms | 0 | Sleep Study | NA | NA |
| Time spent Below 80% SpO2 | 0 | Sleep Study | NA | NA |
| Time spent Below 85% SpO2 | 0 | Sleep Study | NA | NA |
| Time spent below 90% SpO2 | 0 | Sleep Study | NA | NA |
| SAVE_MatlabCustom.csv_sdNNms | 0 | Sleep Study | NA | NA |
| SF36 Body Pain subscale | 1 | General & Mental Health | 1 | NA |
| SF36 General Health subscale | 1 | General & Mental Health | 1 | NA |
| SF36 Trans | 1 | General & Mental Health | 1 | NA |
| SF36 Limit Emotion Subscale | 1 | General & Mental Health | 1 | NA |
| SF36 Limit Physical Subscale | 1 | General & Mental Health | 1 | NA |
| SF36 Mental Subscale | 1 | General & Mental Health | 1 | NA |
| SF36 Physical Subscale | 1 | General & Mental Health | 1 | NA |
| SF36 Social Subscale | 1 | General & Mental Health | 1 | NA |
| SF36 Vitality Subscale | 1 | General & Mental Health | 1 | NA |
| Country | 0 | Demographic | NA | NA |
| Recruitment Site Mnemonic | 0 | Demographic | NA | NA |
| Smoking Status | 1 | Substance Use | 1 | NA |
| Total Number of Snoring Events in Evaluation | 1 | Sleep Study | 1 | NA |
| Duration of Oximetry Analysis | 0 | Sleep Study | NA | NA |
| Duration of Oximetry Analysis | 0 | Sleep Study | NA | NA |
| Subgroup of Age | 0 | NA | NA | NA |
| Subgroup of CV History | 0 | NA | NA | NA |
| Subgroup of Diabetes History/Known at Baseline | 0 | NA | NA | NA |
| Subgroup of ESS | 0 | NA | NA | NA |
| Subgroup based on BMI | 0 | NA | NA | NA |
| Subgroup of ODI | 0 | NA | NA | NA |
| Subgroup of Region (China/Other) | 0 | NA | NA | NA |
| T90 Area | 0 | Sleep Study | NA | NA |
| T90 Area (%) | 1 | Sleep Study | 1 | NA |
| T90 (%) | 1 | Sleep Study | 1 | NA |
| T90 | 0 | Sleep Study | NA | NA |
| Time spent below 80% SpO2 (%) | 1 | Sleep Study | 1 | NA |
| Time spent below 85% SpO2 (%) | 1 | Sleep Study | 1 | NA |
| Time spent below 90% SpO2 (%) | 0 | Sleep Study | NA | NA |
| Total Hypoxic Load | 1 | Sleep Study | 1 | NA |
| Usable sleep time | 1 | Sleep Study | 1 | NA |
| NA | 0 | NA | NA | NA |
| Snoring Loudness | 0 | NA | NA | NA |
| Snorer | 0 | NA | NA | NA |
| How often have Breathing Pauses been noted (code) | 0 | NA | NA | NA |
| Do you Snore? Frequency (code) | 0 | NA | NA | NA |
| Taken Organic Nitrates | 1 | CV Medication | 1 | NA |
| CAD only | 0 | Comorbidity | NA | NA |
| TIA only | 0 | Comorbidity | NA | NA |
| Other form of Cardiovascular Disease | 0 | Comorbidity | NA | NA |
| Diabetes Mellitus | 1 | Comorbidity | 1 | NA |
| Any Heart Disease | 0 | Comorbidity | NA | NA |
| TIA only (no Stroke) | 1 | Comorbidity | 1 | NA |
| Stroke only (no TIA) | 1 | Comorbidity | 1 | NA |
| Both Stroke and TIA | 1 | Comorbidity | 0 | Rare or Invariant |

**Table S2.** European Quality of Life Score, Epworth Sleepiness Scale, Framingham Risk Score, and Average Individual Treatment Effect by Prototype

| **Prototypes** | **Variables** | **Sample Size** | **Mean** | **Standard Error** | **95% Confidence Interval - Lower Bound** | **95% Confidence Interval - Upper Bound** |
| --- | --- | --- | --- | --- | --- | --- |
| 1 | European Quality of Life Score | 586 | 0.72 | 0.014 | 0.69 | 0.75 |
|  | Epworth Sleepiness Scale | 634 | 7.92 | 0.147 | 7.63 | 8.21 |
|  | Framingham Risk Score | 607 | 23.3 | 0.296 | 22.8 | 23.9 |
|  | Average Individual Treatment Effect | 610 | -0.04 | 0.010 | -0.06 | -0.02 |
| 2 | European Quality of Life Score | 414 | 0.83 | 0.012 | 0.81 | 0.86 |
|  | Epworth Sleepiness Scale | 448 | 7.26 | 0.171 | 6.92 | 7.60 |
|  | Framingham Risk Score | 429 | 19.8 | 0.374 | 19.1 | 20.5 |
|  | Average Individual Treatment Effect | 441 | 0.01 | 0.011 | -0.01 | 0.03 |
| 3 | European Quality of Life Score | 402 | 0.90 | 0.012 | 0.87 | 0.92 |
|  | Epworth Sleepiness Scale | 439 | 7.47 | 0.164 | 7.15 | 7.79 |
|  | Framingham Risk Score | 429 | 17.9 | 0.333 | 17.3 | 18.6 |
|  | Average Individual Treatment Effect | 423 | -0.05 | 0.009 | -0.07 | -0.03 |
| 4 | European Quality of Life Score | 476 | 0.85 | 0.011 | 0.82 | 0.87 |
|  | Epworth Sleepiness Scale | 504 | 7.43 | 0.157 | 7.13 | 7.74 |
|  | Framingham Risk Score | 488 | 18.8 | 0.320 | 18.2 | 19.4 |
|  | Average Individual Treatment Effect | 488 | -0.07 | 0.010 | -0.09 | -0.05 |
| 5 | European Quality of Life Score | 593 | 0.90 | 0.009 | 0.88 | 0.92 |
|  | Epworth Sleepiness Scale | 651 | 6.88 | 0.142 | 6.61 | 7.16 |
|  | Framingham Risk Score | 610 | 17.1 | 0.331 | 16.5 | 17.8 |
|  | Average Individual Treatment Effect | 630 | 0.02 | 0.007 | 0.00 | 0.03 |
